# Telehealth Applied to Deliver In-situ Behavioral Skills Training to Reduce Car Seat Misuse During the Covid-19 Pandemic

**DOI:** 10.1101/2020.12.21.20248679

**Authors:** James M. DeCarli

**Author notes:** Correspondence concerning this article should be addressed to James M. DeCarli, 744 So. Windsor Boulevard, Los Angeles, California 90005, United States. The author has no known conflict of interest to disclose.

## Abstract

**Objective:** The use of telehealth has been a common approach to deliver health education before and during the COVID-19 pandemic. However, its ability to apply behavioral skills training (BST) for CRS education has been undocumented. This study assessed the efficacy of telehealth to deliver in-situ behavioral skills training (BST) to teach expectant parents how to install and use their child restraint system (CRS) to reduce misuse and improve retention during the COVID-19 pandemic.

**Method:** A repeated measures group design was used to evaluate 171 individual participants, in a 37-step CRS task analysis for baseline, BST, and follow-up. Performance across all participants was aggregated for each task analysis. Participants were recruited from National Highway Traffic Safety Administration car seat fitting stations during the Covid-19 pandemic between March through July, 2020.

**Results:** Baseline results identified significant critical misuse across participants. With BST, delivered with telehealth, misuse decreased by 97% among 37 task objectives. A 2-week follow-up evaluation concluded that 100% of participants retained the skills they mastered during BST.

**Conclusions:** This study suggests the use of telehealth, as a method of BST delivery for CRS education, is an effective approach to reduce CRS misuse and the burden of child occupant motor vehicle injury. It was found to empower participants and improve their self-confidence, while ensuring the safety of their child occupant. While it was found to be an effective approach for expectant parents during the COVID-19 pandemic, it also has broader child passenger safety program and train-the-trainer implications beyond the COVID-19 pandemic.

---

Motor vehicle occupant injury is one of the leading causes of hospitalizations and fatality for children in the United States.^1^ In 2017, 675 were killed and 116,000 were injured in car crashes.^2^ Injuries among child occupants often result in long term disability.^3 4^ When a child is properly restrained in a child restraint system (CRS), a car seat or booster seat, fatality risk is reduced by 71% for infants, 54% among toddlers and 59% among children in booster seats.^5 6^ Despite child passenger safety laws and educational efforts by Certified Child Passenger Safety Technicians (CPST), critical CRS misuse continues to put child occupants at risk. Misuse is most common among younger children, with 83.9% for infant CRS, 83.5% for rear-facing convertible, 81.9% for forward-facing convertible, and 79.3% for forward-facing only.^7^ Misuse can be attributed to varying designs of CRS’s and motor vehicle seats, methods of installation and misunderstood CRS instructions.^8 9^

Early in 2020, the coronavirus disease-2019 (COVID-19) pandemic posed unprecedented challenges resulting in “Shelter-In-Place” orders, that had the potential to further increase CRS misuse.^10 11^ While CRS education occurs within close quarters inside the parent’s vehicle, this would violate orders and put parents at risk of COVID-19 transmission. As a result, CPST’s were no longer available to assist parents. This put expectant parents and their newborns at risk of CRS misuse. Studies estimate that 96% of parents feel that their CRS is correctly installed, but over 72% are incorrect.^12^ Even during COVID-19, babies were still being born. On average, seven babies are born every 60 seconds in the United States and most are driven home from the hospital.^13^ Estimates have found that 95% of newborn babies are improperly restrained on their first ride home from the hospital.^14^ While misuse was a problem before the pandemic, during it, and with no CPST’s available, this increased the burden of injury even further.

To help fill this gap, telehealth, a virtual remote technology approach, could help expectant parents with their CRS. Telehealth is a well-documented health education approach, applied with a variety of telecommunication platforms, and found to be equivalent to in-person sessions.^15 16^ Telehealth was rarely used in health education before the COVID-19 pandemic. However, during the pandemic, it has become highly recognized and more widely used.^17^

CRS education requires a hands-on, in-person approach, with complicated tasks, that when teaching virtually, could become challenge. However behavioral skills training (BST), a validated method designed to teach new and complex skills, could help meet this challenge.^18-21^ BST is simulated by role playing using four procedures, including; verbal instructions, modeling, rehearsal, and feedback.^22^ During instructions, the educator describes each task until the learner understands what they are expected to perform. Instructions are paired with modeling to improve learning. During modeling the educator demonstrates how the task is to be performed and repeated. During rehearsal, the learner performs each task, while the educator provides feedback. Each task is repeated until the learner has mastered the skill without error. The purpose of this study was to assess the efficacy of using telehealth to deliver in-situ BST to teach parents how to install and use their CRS properly to reduce misuse and improve retention during the Covid-19 pandemic.

## Method

### Participants

Participants included 87 expectant women in their 37^th^ to 39^th^ week of pregnancy and 84 of their partners or family members for a total of 171 participants. Eligibility criteria required having a CRS and motor vehicle. Participants were recruited from telephone inquiries between March through July, 2020. Incoming calls were received from NHTSA inspection stations and 21 birthing hospitals throughout southern California. Of all incoming calls, 94% accepted recruitment into the study. Ages of participants ranged from 32 to 67. Eighty-one percent of participants reported English as their primary language followed by 12% Spanish, and 7% Asian/Islander. Educational level of participants included 57% college/bachelor’s degree or higher, followed by 43% having completed high school. Two participants reported installing a CRS at least once in their lifetime with no prior training.

### Materials and Setting

Each individual training session and assessment consisted of two stages, one inside the participants vehicle and one with their CRS. Sessions were conducted at the participants’ home using telehealth. The telecommunication platform WhatsApp was used to deliver BST for CRS education.^23^ Participants were expectant women and their partner or other family member. Materials of the participant included their mobile device with WhatsApp downloaded, their CRS, a doll or a stuffed animal, and their motor vehicle. For demonstration purposes the educator had an infant CRS and base, with a training doll.

### Dependent Variables and Operational Definitions

The dependent variable was the number of correct skills learned by each participant for installing the CRS in the vehicle and restraining the training doll in the CRS. This included six target behaviors with a 14-step task analysis for inside the vehicle (Table 1) and a 23-step task analysis in the CRS (Table 2) for a total of a 37-step task analysis.

**Table 1.** Stage 1: Target Behavior and Task Objectives Inside the Vehicle (14-step task analysis)

| Target Behavior | Task Objectives |
| --- | --- |
| Occupant Protection:<br>Demonstrate the basic functions and use of restraint systems and occupant protection in your vehicle | Identify safest location for CRS<br>Identify the LATCH system in the vehicle<br>Identify and demonstrate seat belt locking mechanism<br>Identify air bag locations |
| Child Restraint System (CRS):<br>Demonstrate the methods for installing your specific car seat in your vehicle | Identify car seat LATCH system, lower anchors and top tether<br>Identify belt path location for seat belt or LATCH<br>Identify how to level at 30- to 45-degree angle<br>Identify rear-facing and forward-facing recommendations<br>Identify if center buckle is adjustable |
| Installation and testing:<br>Install your car seat in your vehicle in the safest location and test for proper installation | Identify the safest location in the vehicle<br>Level the CRS according instructions of CRS<br>Install CRS or base using appropriate LATCH or Seat belt<br>Test the CRS side-to-side at belt path & back to front less than 1-inch. If rear-facing: a) angle 45-degree, b) distance between back of CRS and back of front vehicle seat for rear-facing, c) other CRS specific manufacturer requirements<br>If infant CRS with base, install CRS without base, if permitted according to manufacturer instructions. Then test, similarly according to rear-facing testing methods |

**Table 2.** Stage 2: Target behavior and task objectives with training doll inside the child restraint system (CRS) [23-step task analysis]

| Target Behavior | Task Objectives |
| --- | --- |
| Occupant protection:<br>Demonstrate basic functions of restraining a child in your CRS according to manufacturer recommendation | Identify how to adjust harness straps to fit child<br>Identify how to adjust center buckle to fit child<br>Identify where the chest clip is to be located<br>Identify how long the newborn insert is to be used<br>Identify specific CRS manufacturer details |
| Training doll:<br>Properly restrain the training doll in your car seat according to your CRS manufacturer recommendations and test for correct use | Demonstrate putting training doll in the CRS<br>Demonstrate adjusting harness straps at or below shoulders (rear-facing) or (at or above shoulders for forward-facing)<br>Demonstrate buckling center buckle<br>Demonstrate buckling chest clip<br>Tighten harness straps upward with thumb and index finger<br>Demonstrate sliding chest clip, pulling slack from the top<br>Demonstrate pulling center strap to tighten harness straps<br>Demonstrate the pinch test on legs and shoulders (thumb and index finger pinching without pulling away from child)<br>Demonstrate chest clip to be level with child's arm pits<br>Repeat of the pinch test on legs and shoulders<br>Describe harness strap adjustment as the child grows<br>Demonstrate adjustment if child's chin is touching chest<br>Demonstrate when to use the newborn insert |
| Use of CRS – duration:<br>Identify how long the newborn baby will be able to use the car seat based on recommendations on the manufacturer labels on the CRS | Identify height limitation from label on the CRS<br>Identify weight limitation from label on the CRS<br>Identify when the child will outgrow the CRS<br>Identify how to register the CRS for recalls<br>Identify expiration of CRS |

Task objectives were based on common misuse errors identified by the National Highway Traffic Safety Administration (NHTSA).^24^ Task objectives included CRS instructions, motor vehicle manufacturer recommendations and best practices by CPST’s. Individual participant performance for each task were measured and recorded by the CPST, who was also trained in applied BST. The CPST recorded the number of correct and incorrect tasks, and number of attempts for each participant. These were based on performance task definitions for the CRS installation and securing the trailing doll (Table 3). To compare performance across all 171 participants, the responses for each participant was calculated into a percentage for each step of the 37-step task analysis for baseline, BST, and follow-up evaluation.

**Table 3.** Performance task definitions for child restraint system (CRS) installation and securing training doll in CRS

| Performance Test | Definition |
| --- | --- |
| Safest Location | Center/appropriateness (<1 inch back of front seats for rear-facing) |
| Angle/Level | CRS or CRS base level according to manufacturer instructions |
| LATCH/Seat Belt | Appropriate method based on CRS use and manufacturer instructions |
| Testing (CRS) | Test CRS installation (side-to-side, back-to-front <1 inch) |
| Harness snugness | Pinch test (Unable to pinch and pull away from child) |
| Harness location | At or below child's shoulders rear-facing/at or above for forward-facing |
| Retainer clip | Level with child's arm pits |
| Testing (doll) | Child's chin is not on chest (then modify), then repeat all of the above |

The percentage of responses across all participants were then aggregated for each step of the 37-step task analysis. A repeated measures group design allowed each individual participant to be exposed to all levels of the independent variable (37-step task objectives). The design helps to improve sensitivity of this study by removing individual participant differences, which allows effective analysis of the task objectives that are directly related to misuse. To assess the integrity of implementation, interobserver agreement scores were calculated by the educator by using scoring sheets at baseline, BST, and follow-up. Scoring categories ensured the delivery of each step of the 37-step task analysis and each BST category (instructions, modeling, rehearsal, and feedback). Agreement scores across all educators for interobserver reliability was 100%.

### Procedure

During recruitment, participants were asked to install their CRS, based on the CRS and motor vehicle instructions. They were provided with an electronic informed consent, demographics form, and a questionnaire of their history of CRS use. To measure the efficacy of telehealth to deliver BST for CRS education, this study used a repeated measures group design for baseline, BST and follow-up. Baseline and BST sessions lasted at least 1.5 hours with 12% of sessions lasting 2 hours. Follow-up evaluation was conducted with telehealth 14 days after the session.

### Baseline

When the participant arrived at their session, they had already installed their CRS in their motor vehicle and practiced restraining a doll or stuffed animal in their CRS restraint. This was requested during phone intake. Participants were assessed by the educator with telehealth. For each correct task 1 was recorded, and for an incorrect task zero was recorded. This process was repeated for each step of the 14-step task analysis for the CRS and for each step of the 23-step task analysis with the training doll.

### Behavioral Skills Training Delivered with Telehealth

Telehealth was used to deliver BST to teach the participant each step of the 37-step task analysis. When BST is performed in-person the CPST demonstrates with the participants’ CRS in their vehicle. Teaching virtually, required each BST procedure to be communicated with clear verbal and visual cues. The 37-step task analysis was divided into 6-chains of sequences of CRS education, including three in the motor vehicle and three in the CRS. Each chain was further divided into smaller sub-sections, while incorporating all four procedures of BST to each sub-section (Figure 1).

**Figure 1.**
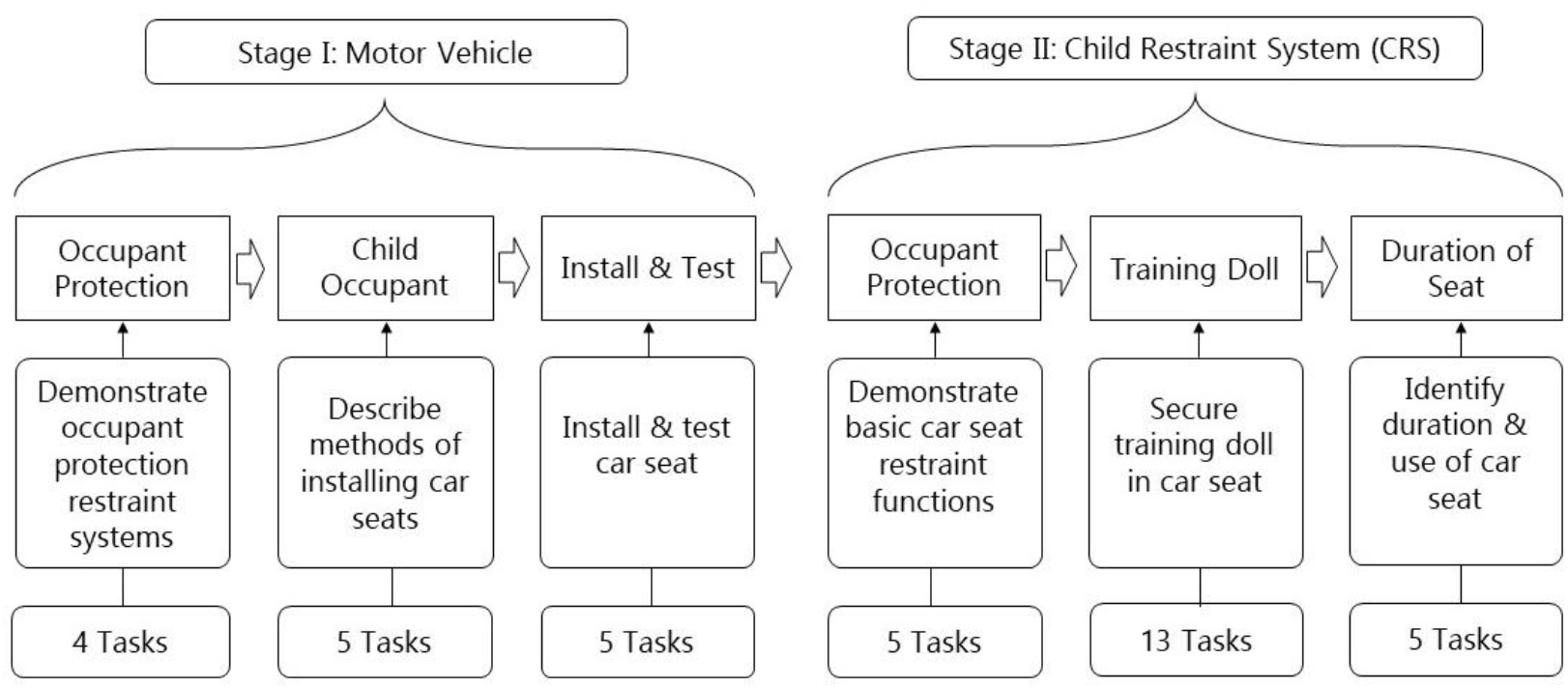
Target Behavior Sequences: A Framework for child restraint system (CRS) Education

Each participant repeated installing their CRS and securing their doll or stuffed animal until they mastered each skill at 100%. Once 100% correct, the participant repeated installing the CRS and securing the training doll between at least 3 times to ensure mastery and not by chance. To control for order effects, counterbalancing modified the order of behavior sequences by randomizing CRS task objectives. Once the participant mastered installing their CRS and securing their training doll at 100% correct, these were repeated three times to improve retention and confidence of correct use. To reduce practice and repetition effects (where a participant could perform better on their second try or exhibit fatigue), once the participant was at 100% correct with the CRS, the educator began another task objective with the same BST process. Once at 100%, the participant would return to the previous task objective and repeat in order to master their skills again at 100%.

### Follow-up

To assess the sustainability of mastery and continued correct CRS use, a follow-up evaluation was conducted with telehealth after 14 days post BST. The participant was instructed to test their CRS, remove and reinstall the CRS without instruction from the educator. This was followed by the participant securing their doll, stuffed animal, or newborn baby into their CRS. The educator observed and assessed the participant performance by recording with the same 37-step task analysis used at baseline and BST sessions.

## Results

The percentage of correct CRS use for the 37-step task analysis, delivered with telehealth, aggregated across all participants during baseline, BST, and follow-up are shown in figure 2 and 3.

**Figure 2.**
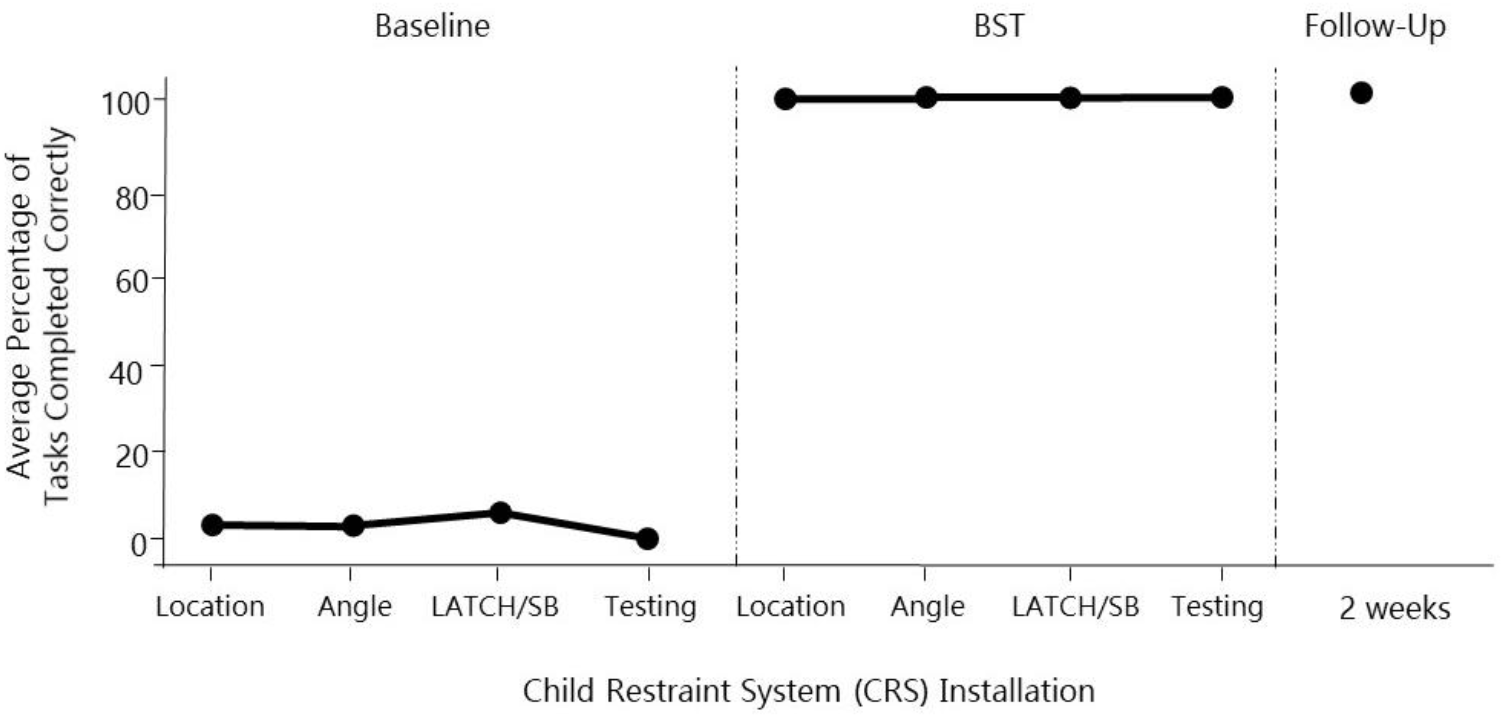
Stage 1: The percentage of correct child restraint system (CRS) installation across all 171 participants during baseline, behavioral skills training (BST), and follow-up delivered with telehealth.

**Figure 3.**
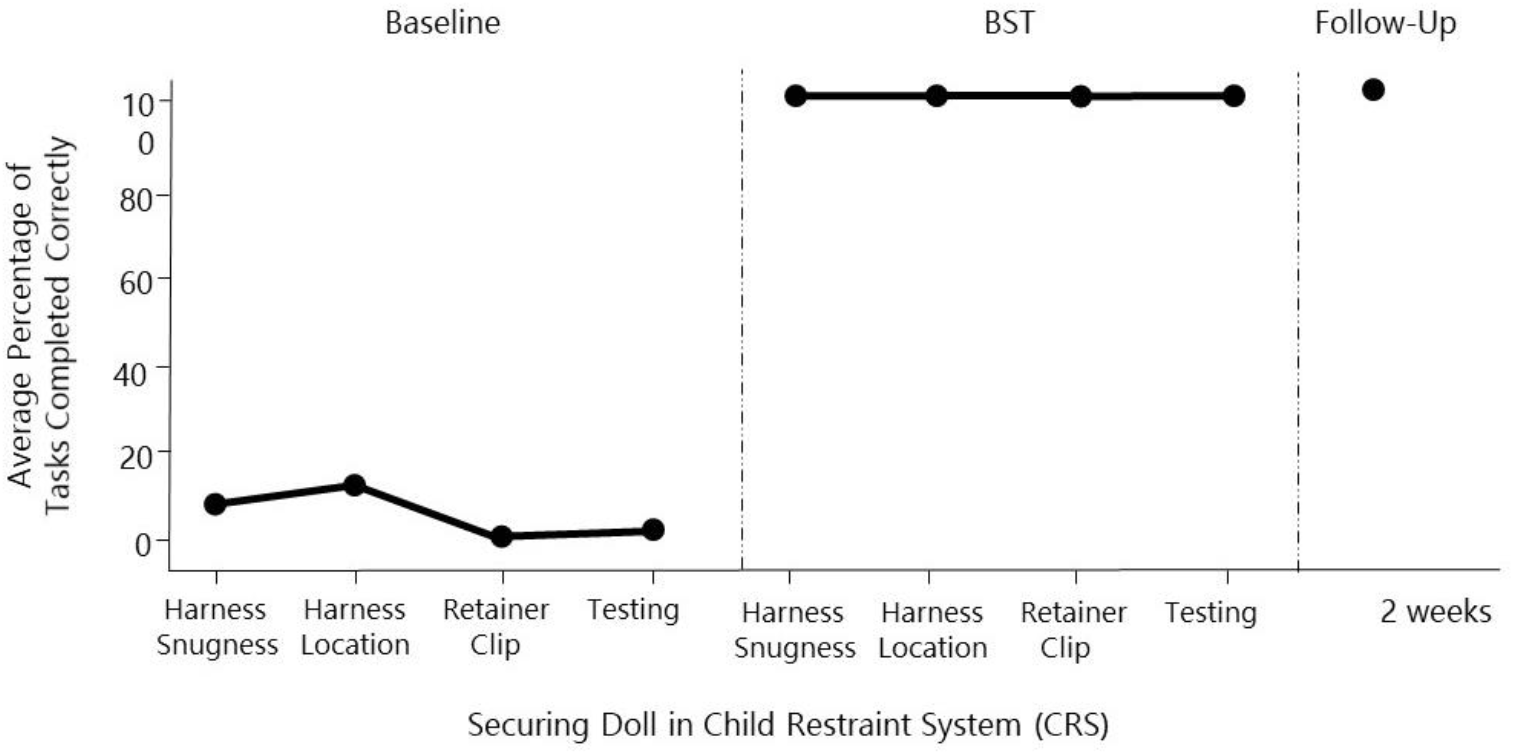
Stage 2: The percentage of restraining of training doll correctly across all 171 participants during baseline, behavioral skills training (BST), and follow-up delivered with telehealth.

During stage-1, the ability to install the CRS correctly across all participants at baseline found a 97% misuse. For stage-2, securing the training doll correctly in the CRS found a 96% misuse. During baseline, common misuse for CRS installation included: location 97%, angle 96%, and LATCH/seat belt 95%. For securing the doll in the CRS misuse included: harness snugness 94%, harness location 91%, retainer clip at 100%, and testing ability 99%. During BST, misuse decreased dramatically with all participants mastering each step of the 37-step task analysis for both the CRS and training doll. At follow-up, it was further observed that the 37 new and complex skills were sustained with no misuse.

## Discussion

This study confirms what is already known in child passenger safety, that misuse is common, especially among new and expectant parents. It adds an important educational tool that helps to reduce CRS misuse and the burden of injury among child occupants. This study is the first to validate the efficacy of a telehealth approach to deliver in-situ BST for CRS education. The results from our study suggest that telehealth is an effective approach in its ability deliver BST to reduce CRS misuse.

The results of our study are consistent with earlier research findings. Baseline results identified common critical CRS misuse.^25-28^ Even though participants were instructed to install their CRS based on CRS and motor vehicle instructions, we observed that instructions and knowledge were not sufficient to provide the complex skills necessary to properly install and use a CRS.^29 30^ We determined that BST procedures provided participants with the hands-on critical skills needed for correct CRS installation and use.^31-33^ While BST was delivered virtually with telehealth, we enhanced verbal and visual cues for each of the four BST procedures. This improved our ability to effectively transfer learning from the educator to participant. This was evident during both BST telehealth and telehealth follow-up, where participant performance resulted in no CRS misuse.

These findings also reflect similar outcomes, before and during the COVID-19 pandemic, which identified the effectiveness of telehealth as a method of BST delivery.^34^ Furthermore, the use of telehealth as a delivery method of BST resulted in outcomes similar to those of in-person sessions.^35-37^ Interestingly, during both telehealth BST and telehealth follow-up, we observed that the virtual approach with BST, placed the responsibility onto the participant. This empowered the participant. It also improved their self-confidence, and removed the common fear of CRS installation.

The 171 participants in this study demonstrated strong external validity. Participants represented expectant parents, in a wide range of demographics from age, language and education. Having BST delivered in-situ, further validated the effectiveness of telehealth as an approach to deliver BST due to the wide range of parent demographics, their lack of CRS experience, and the variety of models of CRS and motor vehicles.

## Limitations

Due to the restrictions from the COVID-19 pandemic, it prevented our ability to have an additional experimenter observe the educator. To compensate, the educator, who was a CPST and trained in BST, recorded interobserver data. To improve efficacy, the scoring sheet was designed, line-by-line for all 37-step task objectives and BST procedures to ensure procedural integrity.

## Conclusion

This study suggests the use of telehealth, as a method of BST delivery for CRS education, is an effective approach to reduce CRS misuse and the burden of motor vehicle injury among child occupants. While it was found to be effective during the COVID-19 pandemic, it is an effective approach to reduce CRS misuse post-pandemic. It also has broader cost-effective child passenger safety and public health program implications that can increase the delivery of injury prevention services to hard-to-reach populations, new parents at hospital discharge, Tribal and rural communities, and geographic locations that have limited or no CPST’s.

## What is already known on this subject

- The misuse of installing and using a child restraint system (car seat or booster seat) is common.
- Car seat manufacturer instructions and motor vehicle occupant recommendations do not result in correct child restraint system (car seat or booster seat)

## What this study adds

- An innovative use of telehealth technology to deliver behavioral skills training as a new and effective tool for Certified Child Passenger Safety Technicians (CPST), beyond the COVID-19 pandemic.
- The use of telehealth to deliver in-situ behavioral skills training (BST) to teach expectant parents how to install and use their car seat will help to reduce misuse.
- Identification on the importance of applied experiential learning (hands-on, practice) in child passenger safety education.
- BST is an effective approach that provides parents with the hands-on, the critical skills needed to properly install and use their car seat.
- Empowers parents and improves self-confidence in child passenger safety.
- A cost-effective approach to improve the quality of car seat education and public health injury prevention programs.
- An approach to deliver, effective car seat education to parents in hard-to-reach populations, new parents at hospital discharge, Tribal and rural communities and locations with limited CPST’s.

## Data Availability

Data included in the manuscript can be made available upon request.

